# Validation of the ACMG/AMP guidelines-based seven-category variant classification system

**DOI:** 10.1101/2023.01.23.23284909

**Authors:** Jian-Min Chen, Emmanuelle Masson, Wen-Bin Zou, Zhuan Liao, Emmanuelle Génin, David N. Cooper, Claude Férec

**Author notes:** **Correspondence:** Jian-Min Chen, MD, PhD, INSERM UMR1078 – EFS – UBO, 22 avenue Camille Desmoulins, 29238 BREST, France.

## Abstract

**Background:** One shortcoming of employing the American College of Medical Genetics and Genomics/Association for Molecular Pathology (ACMG/AMP)-recommended five-category variant classification scheme (“pathogenic”, “likely pathogenic”, “uncertain significance”, “likely benign” and “benign”) in medical genetics lies in the scheme’s inherent inability to deal properly with variants that fall midway between “pathogenic” and “benign”. Employing chronic pancreatitis as a disease model, and focusing on the four most studied chronic pancreatitis-related genes, we recently expanded the five-category ACMG/AMP scheme into a seven-category variant classification system. With the addition of two new classificatory categories, “predisposing” and “likely predisposing”, our seven-category system promises to provide improved classification for the entire spectrum of variants in any disease-causing gene. The applicability and practical utility of our seven-category variant classification system however remains to be demonstrated in other disease/gene contexts, and this has been the aim of the current analysis.

**Results:** We have sought to demonstrate the potential universality of pathological variants that could be ascribed the new variant terminology (‘predisposing’) by trialing it across three Mendelian disease contexts (i.e., autosomal dominant, autosomal recessive and X-linked). To this end, we firstly employed illustrative genes/variants characteristic of these three contexts. On the basis of our own knowledge and expertise, we identified a series of variants that fitted well with our “predisposing” category, including “hypomorphic” variants in the *PKD1* gene and “variants of varying clinical consequence” in the *CFTR* gene. These examples, followed by reasonable extrapolations, enabled us to infer the widespread occurrence of “predisposing” variants in disease-causing genes. Such “predisposing” variants are likely to contribute significantly to the complexity of human genetic disease and may account not only for a considerable proportion of the unexplained cases of monogenic and oligogenic disease but also for much of the “missing heritability” characteristic of complex disease.

**Conclusion:** Employing an evidence-based approach together with reasonable extrapolations, we demonstrate both the applicability and utility of our seven-category variant classification system for disease-causing genes. The recognition of the new “predisposing” category not only has immediate implications for variant detection and interpretation but should also have important consequences for reproductive genetic counseling.

## Background

In 2015, the American College of Medical Genetics and Genomics/Association for Molecular Pathology (ACMG/AMP) recommended a five-category scheme (namely, “pathogenic”, “likely pathogenic”, “uncertain significance”, “likely benign” and “benign”) for classifying variants found in genes underlying classical Mendelian disease entities [1]. This five-category scheme has since become widely used in medical genetics (cited 17904 times according to Google Scholar as of 18 January 2023) despite two fundamental limitations. First and foremost, this scheme was designed specifically to describe variants in genes that cause monogenic Mendelian disorders. In other words, it was never intended to be used to classify all variants in all disease-related genes. Moreover, even in the context of classical Mendelian disease genes, the five-category scheme is inadequate to the task of coping with variants that fall somewhere between “pathogenic” and “benign”. In this regard, it is worth making two points. First, the “likely pathogenic” and “likely benign” categories should not be interpreted as denoting all variants that fall somewhere between “pathogenic” and “benign”; rather, these two terms were proposed by ACMG/AMP to “mean greater than 90% certainty of a variant either being disease-causing or benign to provide laboratories with a common, albeit arbitrary, definition” [1]. Second, ACMG/AMP’s “uncertain significance” category is defined as EITHER when (i) criteria for “pathogenic”, “likely pathogenic”, “benign” and “likely benign” are not met OR (ii) the criteria for benign and pathogenic are contradictory. This ambiguity frequently causes confusion among clinicians, researchers, genetic counselors and database curators alike.

The ACMG/AMP variant classification guidelines have been frequently adapted or refined to suit different contexts (e.g., [2-17]). However, all these adaptations and refinements were made with respect to the specific criteria or rules defining each of the five categories of the ACMG/AMP scheme. The stepwise “ABC system” [18] was considered by its proponents to be an add-on or alternative to the ACMG/AMP guidelines. However, one primary limitation of this system lies in its use of single letter codes (i.e., A through F) for variant classificatory categories, which has made direct comparison with the five-category ACMG/AMP scheme difficult and potentially rather confusing.

Employing chronic pancreatitis as a disease model and focusing on its four most studied genes, we recently extended the five-category ACMG/AMP scheme into a general variant classification framework [19]. In brief, we firstly divided the different genes into “disease-causing” and “disease-predisposing” based upon the integration of multiple layers of evidence with particular focus on their different roles in the pathophysiology of the exocrine pancreas. We then employed two new classificatory categories, “predisposing” and “likely predisposing”, to expand and/or adapt the five-category ACMG/AMP scheme for disease-causing genes and disease-predisposing genes, respectively, leading to the proposition of a two-branch general variant classification framework:

1. A seven-category system (i.e., “pathogenic”, “likely pathogenic”, “predisposing”, “likely predisposing”, “uncertain significance”, “likely benign” and “benign”) for disease-causing genes.
2. A five-category classification system (i.e., “predisposing”, “likely predisposing”, “uncertain significance”, “likely benign” and “benign”) for disease-predisposing genes.

Our two-branch general variant classification framework overcomes the limited scope of the five-category ACMG guidelines since it has the potential to span the entire spectrum of variants in any disease-associated gene. In addition, our general variant classificatory framework has one additional practical advantage: it retains the backbone of the five-category ACMG/AMP scheme, implying that many ACMG/AMP-established rules and criteria for defining the primary variant classificatory categories can be readily adapted or incorporated into our general framework.

The above notwithstanding, both the applicability and utility of our general variant classification framework remain to be demonstrated in other disease/gene contexts. As the first step of this endeavor, we explore herein the applicability and practical utility of the seven-category variant classification system branch by trialing it across three classical Mendelian disease contexts (i.e., autosomal dominant, autosomal recessive and X-linked). Employing a range of illustrative disease/gene/variant examples and reasonable extrapolations, we demonstrate that our seven-category variant classification system has the potential to be widely applicable.

## Results

### Analytical rationale and strategy

Our seven-category variant classification system has two new categories, “predisposing” and “likely predisposing”, by comparison with the five-category ACMG/AMP guidelines [19]. With hindsight, these two new categories may be regarded as being separated from ACMG/AMP’s “uncertain significance” category which, in our view, actually includes two distinct types of variant (Figure 1). The first of these corresponds to those variants for which there is sufficient evidence to support their intermediate status somewhere between “pathogenic” and “benign”, corresponding to our defined *predisposing* category. The second refers to those variants for which there is insufficient evidence for any categorization, and which may be best described as being of “unknown significance”. In the light of this reflection, the “uncertain significance” category previously used in our general variant classification framework was revised here to “unknown significance”. In this regard, it is worth mentioning that both terms, “variant of unknown significance” and “variant of uncertain significance”, have been frequently used in the literature and both are abbreviated as “VUS”.

**Figure 1.**
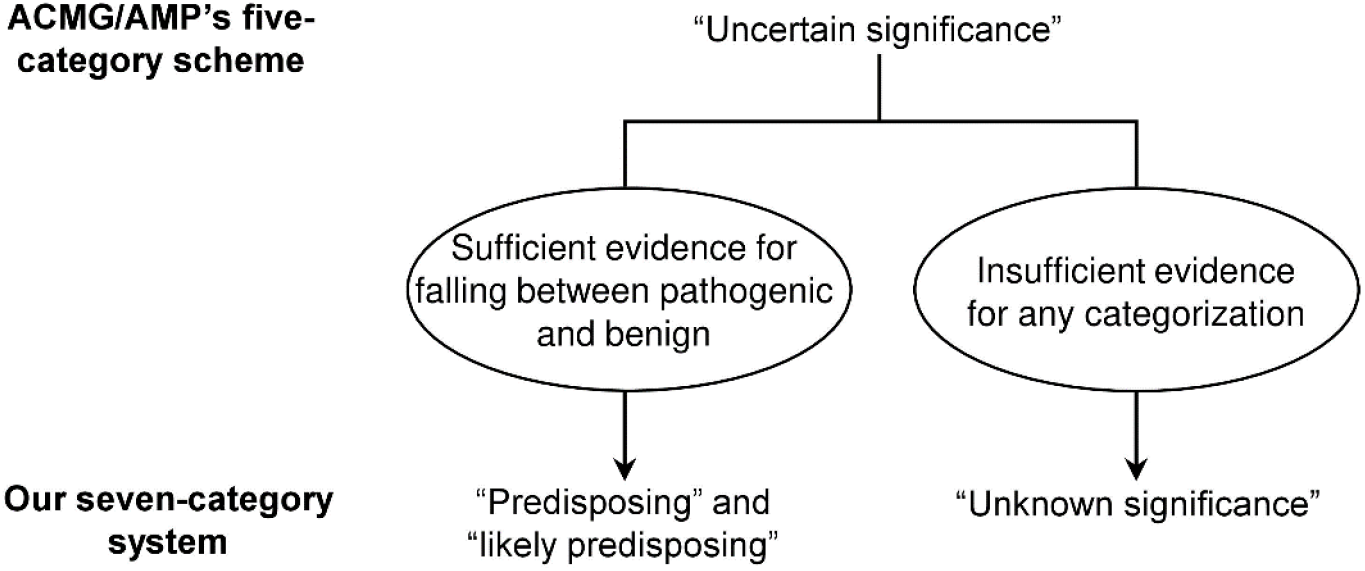
Key differences between our seven-category system and the ACMG/AMP’s five-category scheme. The two new classificatory categories (i.e., “predisposing” and “likely predisposing”) in our seven-category variant classification system were considered here to be separated or split from the ACMG/AMP’s “uncertain significance” category. The “uncertain significance” category previously used in our variant classification framework was also revised here to “unknown significance”.

A comparison of our revised two-branch general variant classification framework with ACMG/AMP’s five-category scheme is provided in Figure 2. The key to demonstrating the applicability and practical utility of our seven-category variant classification system is thus to demonstrate the general occurrence of pathological variants that can be unambiguously (or at least reasonably) allocated to one of the two new categories, “predisposing” and “likely predisposing”.

**Figure 2.**
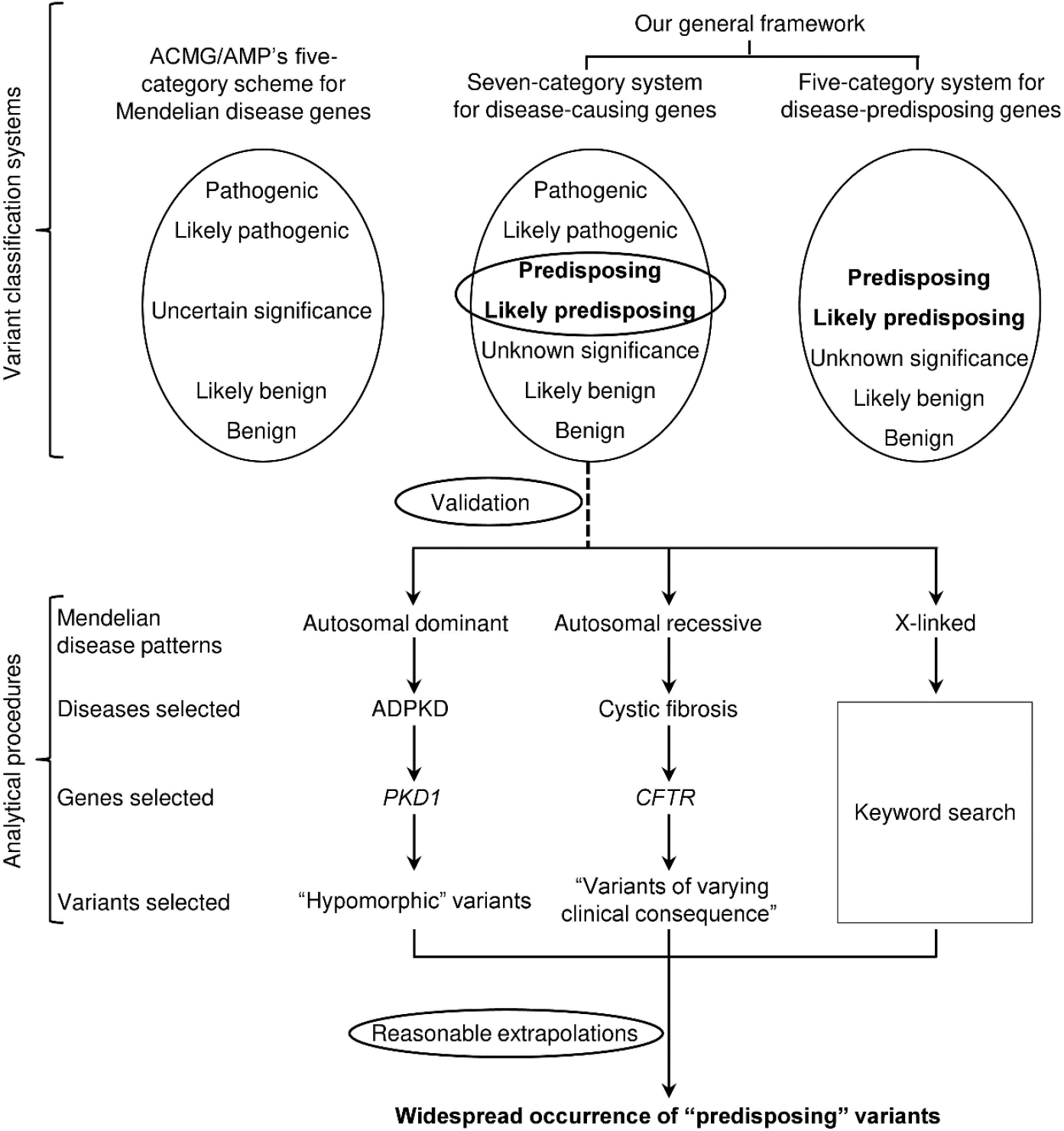
Illustration of the aims and analytical procedures adopted in this study. In the top panel, our revised two-branch general variant classification framework is contrasted with ACMG/AMP’s five-category scheme, with the two new classificatory categories, “predisposing” and “likely predisposing”, being highlighted in bold. The branch validated in the present study, the seven-category variant classification system for disease-causing genes, is connected to the analytical procedures by a dotted line. The validation focuses upon on the two new classificatory categories, which are highlighted in bold and circled by horizontal ovals. ACMG/AMP, the American College of Medical Genetics and Genomics/Association for Molecular Pathology; ADPKD, autosomal dominant polycystic kidney disease. *CFTR*, cystic fibrosis transmembrane conductance regulator; *PKD1*, polycystin-1.

The terms “predisposing” and “likely predisposing” were proposed by analogy to ACMG/AMP’s terms “pathogenic” and “likely pathogenic” as well as “benign” and “likely benign”. As such, “likely predisposing” should denote a greater than 90% certainty of a variant being predisposing. For the purpose of establishing core principles, we shall not attempt to distinguish the two new categories, “predisposing” and “likely predisposing”, in the current analysis.

The “predisposing” category was only recently proposed by us [19]. No attempt had been made in previous studies to allocate variants into a “predisposing” category although the descriptor “predisposing” has been frequently (and generally rather loosely) used in the literature to describe pathologically significant variants. Faced with this situation, as well as the unrealistic prospect of being able to analyze all relevant variants in all disease genes, we opted to adopt a simple practical approach. As mentioned above, our proposed general variant classification framework was built upon sound data from a single model disease, chronic pancreatitis, and its four most extensively studied genes [19]. In similar vein, we shall herein attempt to build our case using illustrative genes/variants causing respectively autosomal dominant, autosomal recessive and X-linked diseases. Findings from these illustrative examples will finally be generalized so as to be applicable to all disease-causing genes.

Before delving into the details of our analysis (Figure 2), several points are worth noting. First, we initiated our analysis with variants in two genes underlying respectively an autosomal dominant disease and an autosomal recessive disease, in both of which we have expertise [20-25]. Findings from these examples were then used to inform keyword searches in the context of X-linked diseases. We would like to reiterate that these analytical procedures were intended to identify some but not all pertinent examples. Second, pathologically relevant variants in the primary chronic pancreatitis-causing gene, *PRSS1* (OMIM #276000; encoding cationic trypsinogen), which originally prompted us to propose the seven-category variant classification system, give rise to a gain of trypsin function or a gain of proteotoxicity [19]. By contrast, the pathologically relevant variants that will be discussed below lead to the functional loss of their affected genes/proteins. In these disease genes, predicted loss-of-function (i.e., nonsense, frameshift, large genomic deletion, and obligate splice site) variants have almost invariably been classified as “pathogenic” whereas variants in regulatory or deep intronic regions have often been classified as “benign” or “likely benign”. By contrast, it is often difficult to confidently classify missense variants appropriately without supporting functional data. Thus, not surprisingly, the “predisposing” variants to be described below involve predominantly missense variants. Third, “hypomorphic” (partial but not complete loss of function) variants in the genes to be discussed below may also be considered to be “predisposing”. However, “hypomorphic” variants in some other gene/disease contexts may either be “pathogenic” or “benign” (a simple analogy may be that while a complete loss-of-function variant in one gene may be harmless, in another gene it is disease-causing). Moreover, to the best of our knowledge, there is no consensus with respect to a threshold of partial functional loss as a means to define “hypomorphic”. Fourth, we have not performed any *in silico* predictions to assist the classification of missense variants owing to the relatively poor overall performance of existing prediction tools in relation to this type of variant [26]. Moreover, the functional effects of “predisposing” missense variants are in principle more refractory to prediction than “pathogenic” missense variants (e.g., [27]). Lastly, selected variants will be evaluated with respect to their classifications in ClinVar [28] and respective authoritative disease/gene-specific databases, with a view to illustrating the dilemmas frequently encountered in classifying this type of variant.

### Autosomal dominant diseases

We selected autosomal dominant polycystic kidney disease (ADPKD) as a model of an archetypal autosomal dominant disease. *PKD1* (OMIM #601313), which encodes a 4303 amino acid receptor-like transmembrane protein, polycystin-1, is the primary causative gene for ADPKD located on chromosome 16p13.3 [29, 30]. It has been subjected to extensive study since its identification nearly 30 years ago [31], with 2322 different variants being listed in the ADPKD Variant Database [32]. Of particular relevance, all 815 truncating variants (i.e., frameshift, nonsense, obligate splice site and large rearrangements) were classified by the ADPKD Variant Database as “pathogenic”. By contrast, of the 782 missense variants, 277 have been classified as “likely pathogenic”, 203 have been classified as “VUS” and 302 have been classified as “likely benign”. In other words, none of the 782 missense variants have been classified as either “pathogenic” or “benign”; a reflection of the dire state of affairs in which, as yet, no functional assay has been made available to ascertain the impact of *PKD1* missense variants.

Whilst *PKD1* haploinsufficiency due to rare heterozygous variants gives rise to ADPKD, the inheritance of two ADPKD-causing *PKD1* alleles is a cause of recurrent fetal loss [33, 34]. Consistent with this latter observation, *Pkd1* knockout mice are not viable embryonically [35]. However, mice homozygous for specific “hypomorphic” *Pkd1* alleles developed polycystic kidney disease (PKD) and were viable [36, 37]. Interestingly, human “hypomorphic” *PKD1* variants have also been reported in the literature, in both a homozygous and compound heterozygous context. Such variants reported up until the middle of 2014 [38-42], which were previously reviewed by us [22], will be used here as illustrative examples (Table 1). Specifically, all three individuals homozygous for *PKD1* hypomorphic variants presented with PKD whereas the corresponding heterozygotes had no or only a few renal cysts. As far as the compound heterozygotes were concerned, two situations were evident. In the first, both variants were missense, with all single heterozygotes being unaffected whilst all compound heterozygotes presented with PKD. In the second situation, the compound heterozygotes comprised a missense variant and an unambiguous pathogenic variant; all missense variant heterozygotes were unaffected but all compound heterozygotes were characterized by severe PKD. Presumably, these “hypomorphic” missense variants caused a partial functional loss of the affected allele, albeit a loss that, in simple heterozygosity, did not result in the threshold of PKD causation being breached. However, these variants, when inherited either in the homozygous or compound heterozygous state, reduced *PKD1* function to a level below that of the disease-causing threshold. One such missense variant, p.Arg3277Cys (detected once as a homozygote and twice as a compound heterozygote (Table 1)), was studied in a knock-in mouse model. The homozygous knock-in mice were estimated to exhibit ∼40% of the Pkd1 function of wild-type mice [43]. This suggests that a simple p.Arg3277Cys heterozygote (not PKD-causing) would be expected to retain 70% of normal PKD1 function (i.e. 20% higher than the 50% PKD1 function in heterozygotes carrying a typical “pathogenic” *PKD1* variant).

**Table 1.** Illustrative examples of “hypomorphic” *PKD1* variants*

| Variant(s)<sup>a</sup> | Reference | Classification |  |
| --- | --- | --- | --- |
|  |  | ADPKD variant database | ClinVar |
| **Homozygote** |  |  |  |
| c.[3133G>A;4709C>T]<br>(p.[Val1045Met;Thr1570Met]) | [38] | p.Val1045Met, VUS;<br>p.Thr1570Met, VUS | p.Val1045Met, NA;<br>p.Thr1570Met, CIP (P × 1, VUS × 1) |
| c.9563A>G (p.Asn3188Ser) | [39] | p.Asn3188Ser, VUS | p.Asn3188Ser, NA |
| c.9829C>T (p.Arg3277Cys) | [39] | p.Arg3277Cys, VUS | p.Arg3277Cys, CIP (P × 4, LP × 2, VUS × 2) |
| **Compound heterozygote** |  |  |  |
| c.[**3312dupC**];[6749C>T]<br>(p.[**Val1105fs**];[Thr2250Met]) | [40] | p.Thr2250Met, likely benign | p.Thr2250Met, CIP (VUS × 2, B × 1, LB × 1) |
| c.[3820G>A];[3820G>A;8716G>A]<br>(p.[Val1274Met];[Val1274Met;Gly2906Ser]) | [41] | p.Val1274Met, VUS;<br>p.Gly2906Ser, likely benign | p.Val1274Met, NA;<br>p.Gly2906Ser, VUS (single submitter) |
| c.[**4199delT**];[12413G>A]<br>(p.[**Leu1400fs**];[Arg4138His]) | [41] | p.Arg4138His, VUS | p.Arg4138His, NA |
| c.[**6472C>T**];[9829C>T]<br>(p.[**Gln2158\***];[Arg3277Cys]) | [39] | p.Arg3277Cys, VUS | p.Arg3277Cys, CIP (P × 4, LP × 2, VUS × 2) |
| c.[6658C>T];[9829C>T]<br>(p.[Arg2220Trp];[Arg3277Cys]) | [38] | p.Arg2220Trp, VUS;<br>p.Arg3277Cys, VUS | p.Arg2220Trp, VUS (single submitter)<br>p.Arg3277Cys, CIP (P × 4, LP × 2, VUS × 2) |
| c.[**7915_7916ins20**];[8293C>T]<br>(p.[**Arg2639fs**];[Arg2765Cys]) | [39] | p.Arg2765Cys, VUS | p.Arg2765Cys, CIP (VUS × 3, B × 2, LB × 1) |
| c.[**8259C>G**];[6763C>T]<br>(p.[**Tyr2753\***];[Arg2255Cys]) | [41] | p.Arg2255Cys, VUS | p.Arg2255Cys, NA |
| c.[8293C>T];[9313C>T]<br>(p.[Arg2765Cys];[Arg3105Trp]) | [39] | p.Arg2765Cys, VUS;<br>p.Arg3105Trp, VUS | p.Arg2765Cys, CIP (VUS × 3, B × 2, LB × 1);<br>p.Arg3105Trp, VUS (VUS × 2) |
| c.[**8362_8363ins34**];[5848G>A]<br>(p.[**Ser2788fs**];[p.Val1950Met]) | [42] | p.Val1950Met, VUS | p.Val1950Met, VUS (single submitter) |
| c.[**11457C>A**];[8293C>T]<br>(p.[**Tyr3819\***];[Arg2765Cys]) | [39] | p.Arg2765Cys, VUS | p.Arg2765Cys, CIP (VUS × 3, B × 2, LB × 1) |

ADPKD, autosomal dominant polycystic kidney disease; B, benign; CIP, conflicting interpretations of pathogenicity; LB, likely benign; LP, likely pathogenic;
NA, not available; P, pathogenic; VUS, variant of uncertain (unknown) significance.
\*Variants previously reviewed in [22].
<sup>a</sup>Definite pathogenic variants are highlighted in bold.

Keyword searches in PubMed using “*PKD1*” plus “homozygote”, “incomplete penetrant”, “hypomorphic”, “biallelic” or “trans heterozygous” identified further studies reporting *PKD1* “hypomorphic” variants [44-49]. Notably, the Durkie study reported a high prevalence of biallelic inherited “hypomorphic” *PKD1* variants in very early onset PKD [48]. Of particular relevance, Durkie and colleagues discussed the various difficulties encountered in classifying this particular type of *PKD1* variant. First, they considered that it was challenging to use the current ACMG/AMP variant classification guidelines to classify *PKD1* “hypomorphic” variants as these variants “do not function as classic loss-of-function variants causing autosomal dominant disease” [48]. Moreover, they also regarded the additional terms (i.e., “reduced penetrance” and “ultralow penetrance”) used in the literature to describe this kind of variant [50] as confusing. Further, they considered the ClinGen Consortium Low Penetrance/Risk Allele Working Group-recommended terminology for describing reduced penetrance variants, namely the use of the ACMG/AMP categories plus a quantitative descriptor (i.e., “likely pathogenic, low penetrance” or “likely pathogenic, reduced penetrance”), to be inappropriate for describing *PKD1* “hypomorphic” variants; “the scenario for *PKD1* hypomorphic variants that cause cysts only when inherited in *trans* with another pathogenic or hypomorphic variant does not fall under this definition” [48].

Facing the abovementioned difficulties, Durkie and colleagues opted to use “3H” (the 3 refers to ACMG/AMP’s “uncertain significance” category whilst H denotes “hypomorphic”) for the classification of the “hypomorphic” *PKD1* variants [48]. Finally, Durkie et al. noted that hypomorphic variants are not unique to *PKD1* and cited several additional disease/gene examples with this type of variant (viz. maturity-onset diabetes of the young (MODY)/*HNF1A* [51], *RFX6* [52] and *HNF4A* [53]; Parkinson’s disease/*VPS35* [54] and *LRRK2* [55]; retinal dystrophy/*ABCA4* [56]; and Joubert syndrome/*SUFU* [57]).

As shown in Table 1, the illustrative examples of “hypomorphic” *PKD1* variants were often classified as “VUS” in the ADPKD variant database [32] and as “VUS” or “conflicting interpretations of pathogenicity (CIP)” in ClinVar [28]. Our proposed “predisposing” category provides a perfect fit for the “hypomorphic” *PKD1* variants. Intuitively, findings from the “hypomorphic” *PKD1* variants can be generalized to all autosomal dominant diseases owing to haploinsufficiency (Figure 3).

**Figure 3.**
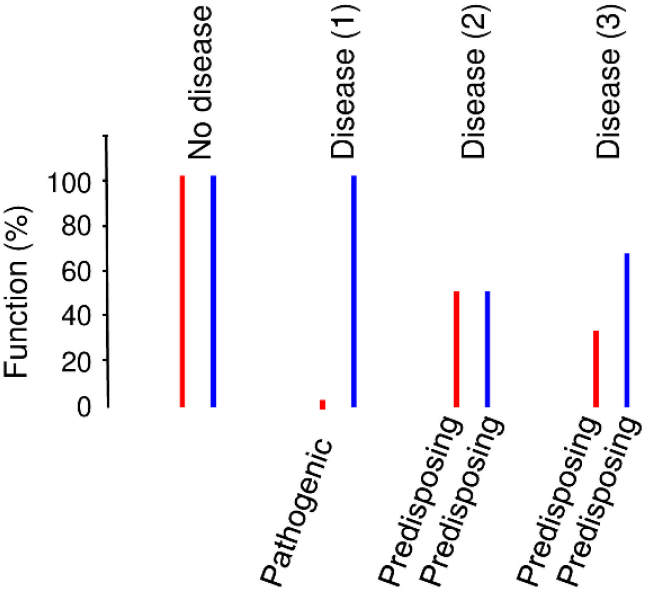
“Predisposing” variants in autosomal dominant diseases caused by haploinsufficiency. This model was generalized from the so-called “hypomorphic” *PKD1* variants. Disease (1), disease caused by a typical “pathogenic” variant leading to the complete or almost complete loss of the affected allele. Disease (2) and disease (3) represent two situations in which the disease is caused by a combination of two “predisposing” variants.

There is one final point to make. In classical Mendelian diseases, disease prevalence has been employed to estimate a maximum allele frequency threshold for a “pathogenic’ variant (e.g., [58-62]). The prevalence of ADPKD in Europe and USA is no higher than 5 in 10,000 inhabitants [63, 64]. This means that a typical *PKD1* “pathogenic” variant (leading to the complete or almost complete loss of the affected allele) cannot exceed an allele frequency of 0.00025 in the corresponding normal population. However, a *PKD1* “hypomorphic” variant may exceed such a maximal allele frequency threshold owing to the absence of an associated clinical phenotype in the heterozygous state [48]. For example, the highest sub-population allele frequency (i.e., 0.0004532 [65]) of the aforementioned *PKD1* p.Arg3277Cys variant was found in the non-Finnish European population. This is ∼1.8 fold higher than the estimated maximum allele frequency of 0.00025 for a typical *PKD1* “pathogenic” variant in Europeans. In short, some pathologically relevant “hypomorphic” variants are likely to have been inadvertently filtered out using the maximal allele frequency threshold established for a typical “pathogenic’ variant.

### Autosomal recessive diseases

We selected cystic fibrosis (CF) [66] as an archetypal autosomal recessive disease. *CFTR* (cystic fibrosis transmembrane conductance regulator; OMIM #602421) is the sole causative gene for CF [67]. The gene is located on chromosome 7q31.2, contains 27 exons and encodes a 1480 amino acid protein. Whereas the Cystic Fibrosis Mutation Database (CFTR1) [68] lists 2110 *CFTR* variants reported until Apr 25, 2011, the Clinical and Functional TRanslation of CFTR (CFTR2) database lists 485 variants with detailed genotype-phenotype information gathered from CF patient registries around the world [69] (CFTR2 is led by a team of world-renowned researchers and clinicians, who are seeking to provide expert-reviewed functional and clinical information on *CFTR* mutations). These 485 variants were classified by CFTR2 into four categories, with the category definitions and number of variants in each category being provided in Table 2. CFTR2 uses the term “varying clinical consequence (VCC)” to describe 49 variants that were deemed not to correspond to any of the five ACMG/AMP-defined variant categories; “If CF is the defined phenotype for which ACMG-AMP guidelines are being applied, it might be difficult to assign these variants as pathogenic or likely pathogenic (without clear evidence of CF in some people), but the term VUS is not appropriate because, in many cases, clinical significance is not uncertain” [27].

**Table 2.** Summary of the 485 CFTR2-annotated *CFTR* variants

| Classification | Definition | Number of variants |
| --- | --- | --- |
| CF-causing | The variant in question “causes CF when combined with another CF-causing variant”. | 401 |
| Varying clinical consequence | The variant in question “has varying consequences. Some patients with the variant, combined with another CF-causing variant, have CF. Other patients with the variant, combined with another CF causing variant, do not have CF”. | 49 |
| Non CF-causing | The variant in question “DOES NOT cause CF when combined with another CF-causing variant”. | 24 |
| Unknown significance | The variant in question “is still under evaluation. At this time, the CFTR2 team does not have enough information to determine whether or not it causes CF”. | 11 |

CF, cystic fibrosis; *CFTR*, cystic fibrosis transmembrane conductance regulator; CFTR2, the Clinical and Functional TRanslation of CFTR database.

Importantly, a comprehensive functional assay of 48 selected *CFTR* missense variants with variable expressivity of CF in a native isogenic context revealed that CF-causing (n = 24), VCC (n = 16) and non-CF-causing (n = 6) missense variants displayed <10%, 10-75% and >75% of wild-type CFTR function, respectively. The simplest biological mechanism responsible for the pathological impact of these VCC variants would involve a reduction in expression of the affected *CFTR* alleles so as to reach the CF-causing threshold level. This could come about if these variants (i) were combined with variants that served to reduce *CFTR* expression in modifier genes, (ii) interacted with environmental factors or (iii) were *in cis* with an as yet unidentified functional variant that served to further reduce *CFTR* expression [27, 70]. With regard to the latter, it is important to emphasize that even in the rare instances of reports describing “complete *CFTR* gene sequencing” [71, 72], variants identified in deep intronic and remote regulatory regions have not been subjected to thorough evaluation.

Of the 16 functionally characterized VCC variants, nine have been shown to retain 10% to 25% of wild-type CFTR function [27]. These nine variants are employed here as illustrative examples with respect to ClinVar classifications. All these variants, with the exception of p.Asp110Glu, were classified as having CIP (Table 3).

**Table 3.** Illustrative examples of CFTR2-annotated variants with “varying clinical consequence”*

| Variant |  | ClinVar classification |
| --- | --- | --- |
| cDNA name | Protein name |  |
| c.14C>T | p.Pro5Leu | CIP (P × 3; LP × 6; VUS × 2) |
| c.330C>A | p.Asp110Glu | Drug response (P × 5; LP × 2; VUS × 1; drug response × 1) |
| c.794T>G | p.Met265Arg | CIP (P × 1; LP × 3; VUS × 2) |
| c.1724T>A | p.Phe575Tyr | CIP (P × 1; LP × 1; VUS × 6) |
| c.1865G>A | p.Gly622Asp | CIP (P × 9; LP × 7; VUS × 2; B × 1) |
| c.3095A>G | p.Tyr1032Cys | CIP (P × 6; LP × 6; VUS × 1) |
| c.3297C>A | p.Phe1099Leu | CIP (P × 2); LP × 1; VUS × 6) |
| c.3458T>A | p.Val1153Glu | CIP (P × 1; VUS × 3) |
| c.3737C>T | p.Thr1246Ile | CIP (P × 5; LP × 2; VUS × 2) |

B, benign; CIP, conflicting interpretations of pathogenicity; LP, likely pathogenic; P, pathogenic; VUS, variant of uncertain (unknown) significance.
\*These variants are characterized by >10% but <25% of wild-type CFTR function [27].

Again, our proposed “predisposing” category would offer a perfect fit for the CFTR2-defined VCC variants. Findings from the illustrative *CFTR*/CF studies were generalized into classical autosomal recessive diseases that were caused by loss-of-function variants (Figure 4).

**Figure 4.**
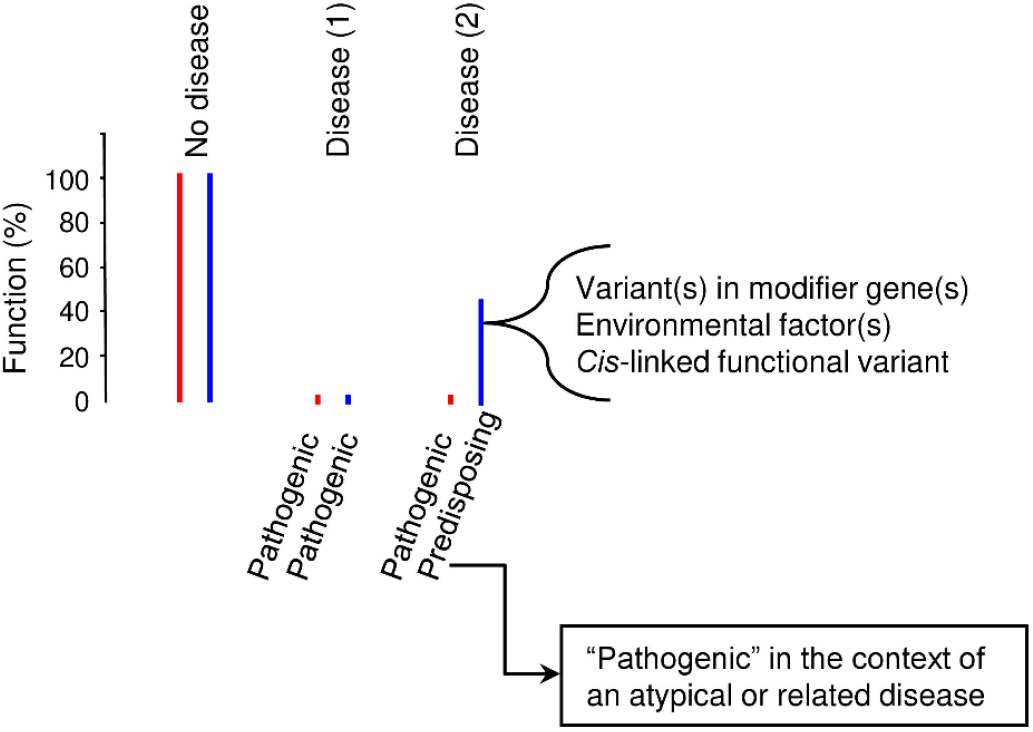
“Predisposing” variants in autosomal recessive diseases caused by loss-of-function variants. This model was generalized from *CFTR* variants with “varying clinical consequence” related to autosomal recessive cystic fibrosis. In disease situation 1, the disease manifests as typical autosomal recessive inheritance. In disease situation 2, the “predisposing” variant must interact with additional variant(s) in modifier genes and/or with environmental factors in order for the disease to be expressed clinically. Alternatively, an as yet unidentified variant may exist in *cis* that serves to further reduce the function of the “predisposing” allele. The curved arrow points to a situation in which the “predisposing” variant can be “pathogenic” for an atypical or related milder disease.

The *CFTR* VCC variants that displayed 10-75% of wild-type CFTR function may be termed “hypomorphic” by comparison to the CF-causing and non-CF-causing variants. Whilst these VCC variants were “predisposing” with respect to CF, they may be “pathogenic” with respect to CFTR-related disorders (clinical entities with features of CF and evidence for the presence of CFTR dysfunction but not meeting the full criteria for diagnosis of CF) [73]; this may be one reason for the often encountered “conflicting interpretations of pathogenicity” in ClinVar with respect to the *CFTR* VCC variants. This possibility is also illustrated in Figure 4.

### X-linked diseases

Whereas females have paired X chromosomes, males possess only a single X chromosome. Moreover, in females, the possession of two X chromosomes leads to a dosage issue which is compensated for by X-inactivation [74, 75]. These two unique properties often blur the standard definitions of dominance and recessiveness in X-linked diseases. Indeed, it has even been recommended that “use of the terms X-linked recessive and dominant be discontinued, and that all such disorders be simply described as following “X-linked” inheritance” [76].

Mindful of the difficulties encountered in analyzing X-linked genes/variants, we sought to use the findings from the abovementioned *PKD1* variants to guide our analysis in the context of X-linked dominant diseases. We began our analysis with X-linked dominant diseases in females, which can be analogous to autosomal dominant diseases. Given that the abovementioned “predisposing” *PKD1* variants were found in either a homozygous or compound heterozygous state, we postulated that homozygous or compound heterozygous variants found in females with a seemingly typical X-linked dominant disease have a high probability of falling into the “predisposin*g*” category. More specifically, the vast majority of female patients with an X-linked dominant condition will be heterozygotes whereas homozygous or compound heterozygous individuals would generally be extremely rare. Therefore, when a homozygous or compound heterozygous individual is encountered, each allele is likely to be hypomorphic because on their own they are insufficient to result in a clinical phenotype. To identify such variants, we performed a keyword search in PubMed using “X-linked dominant” plus “homozygote”, “homozygous”, “hypomorphic”, “compound heterozygote” or “biallelic”. We identified a single candidate homozygous variant that was originally reported in a 30-month-old Indian girl with suspected Rett syndrome (RS) [77]), which will be discussed in detail below.

RS is a severe early onset neurological condition that occurs almost exclusively in females. It is usually inherited in an X-linked dominant manner and is primarily caused by *de novo* heterozygous mutations in the *MECP2* gene [78-80]. Males with RS-causing *MECP2* mutations have severe neonatal encephalopathy that is usually lethal whilst those with non-RS-causing *MECP2* mutations can demonstrate a wide variety of neurological disorders [81]. In the light of findings from the *PKD1*/ADPKD system, some of these non-RS-causing *MECP2* variants may be assumed to represent “hypomorphic” (partial but not complete functional loss) variants as compared to the typical RS-causing *MECP2* variants (complete or almost complete functional loss). Further, some “hypomorphic” variants may be clinically asymptomatic in heterozygous females but cause disease in the homozygous or compound heterozygous state.

The above-mentioned homozygous *MECP2* variant identified in a girl with suspected RS was c.1160C>T (p.Pro387Leu) according to NM_004992.4 or c.1196C>T (p.Pro399Leu) according to NM_001110792.2. Her mother did not carry this variant nor any other mutation in the *MECP2* gene. Her father, who carried the *MECP2* p.Pro399Leu variant, was reported not to exhibit any indication of intellectual disability [77]. The *MECP2* p.Pro399Leu variant had been reported in two studies prior to the Bhanushali report [77]. In the first, the variant was identified in a male patient with sporadic intellectual disability [82]. In the second study, the variant (zygosity not reported but presumably heterozygous) was identified in a female RS patient; it was however deemed to have no pathological relevance owing to its presence in the healthy father [83].

Herein, we further surveyed the *MECP2* p.Pro399Leu variant in ClinVar [28]. Only in one of the seven submissions of this variant was the affected status indicated as “yes” (accession SCV000187889.2). In this single “yes” case, the disorder of the corresponding male patient was diagnosed as “mental retardation, X-linked, syndromic 13” with the *MECP2* variant being of maternal origin. In short, of the aforementioned four male carriers of the *MECP2* p.Pro399Leu variant, two (50%) were reported to exhibit intellectual disability. These limited data may nevertheless be consistent with *MECP2* p.Pro399Leu being a pathologically relevant but “hypomorphic” variant, taking into account the observation that this variant (i) is very rare in the combined gnomAD (the Genome Aggregation Database) male subjects (carrier/allele frequency, 0.00009947 [65]) and (ii) was identified in a gene known to be responsible for various neurological conditions.

In ClinVar [28], the *MECP2* p.Pro399Leu variant was classified as “benign”, “likely benign” and “uncertain significance” by two, three and two submitters, respectively; collectively, it was classified as “benign”. Some submitters provided detailed evidence supporting their classification, with two examples being cited as below;

➢ *Benign, accession SCV002047342*.*1*. “The allele frequency of the p.Pro387Leu (NM_004992.3) variant in *MECP2* is 0.026% in South Asian sub-population in gnomAD, which is high enough to be classified as likely benign based on thresholds defined by the ClinGen Rett/Angelman-like Expert Panel for Rett/AS-like conditions (BS1)” — ClinGen Rett and Angelman-like Disorders Variant Curation Expert Panel.
➢ *Likely benign, accession SCV000698534*.*2*. “This variant was found in 17/186675 control chromosomes (gnomAD) at a frequency of 0.0000911, which is approximately 11 times the estimated maximum expected allele frequency of a pathogenic *MECP2* variant (0.0000083), suggesting this variant is likely a benign polymorphism.” — Women’s Health and Genetics/Laboratory Corporation of America, LabCorp.

The above two submitters classified the *MECP2* p.Pro399Leu variant as “benign” or “likely benign” based primarily upon the maximal disease allele frequency threshold that was estimated for a “pathogenic” *MECP2* variant with respect to typical RS. As with the case of the previously discussed “hypomorphic” *PKD1* p.Arg3277Cys variant, this practice was clearly inappropriate for classifying the “hypomorphic” *MECP2* p.Pro399Leu variant. To better understand this issue, we re-evaluated the prevalence of typical RS and the allele frequency of *MECP2* p.Pro399Leu in gnomAD populations. In this regard, it is important to emphasize that whereas hemizygous *MECP2* p.Pro399Leu males may exhibit intellectual disability, heterozygous *MECP2* p.Pro399Leu females appear almost invariably to be healthy. Thus, to be maximally relevant to our thesis, we re-evaluated these data only in the context of females. The prevalence of classical RS was, at most, one in 10,000 females [84], which corresponds to a maximum allele frequency of 0.00005 for a typical RS-causing *MECP2* variant. The allele frequency of *MECP2* p.Pro399Leu in gnomAD-analyzed females was 0.00006474 [65], which is ∼1.3-fold higher than 0.00005 (it should be remembered that the corresponding ratio for the “hypomorphic” *PKD1* p.Arg3277Cys variant was ∼1.8). Moreover, it should be emphasized that no *MECP2* p.Pro399Leu homozygotes were found in the 61,785 gnomAD-derived females [65].

Taken together, it is not unreasonable to conclude that *MECP2* p.Pro399Leu represents a “hypomorphic” variant that gives rise to intellectual disability in males (with 50% penetrance) in the hemizygous state whilst causing RS in females only in the homozygous state. Bearing this in mind, we may speculate that the previously reported female RS patient carrying a heterozygous *MECP2* p.Pro399Leu [83] may harbor an as yet unidentified “hypomorphic” or “predisposing” variant in the other *MECP2* allele.

Examples of the RS-causing and non-RS-causing variants in the *MECP2* gene may logically imply similar situations in some other X-linked genes, in which heterozygous pathogenic variants (causing complete or almost complete functional loss of the affected allele) in females cause typical X-linked dominant diseases whilst hemizygous “hypomorphic” variants in males (complete or almost complete functional loss is lethal to males) cause a related phenotype; such “hypomorphic” variants may not be associated with a clinical phenotype when found in females in the heterozygous state but will cause disease in females when found in homozygosity or compound heterozygosity. We performed a keyword search in PubMed using “X-linked male female hypomorphic”, identifying multiple X-linked genes that appear to be compatible with the above situation (e.g., *EBP* [85], *NAA10* [86], *BCOR* [87], *NSDHL* [88], *IKBKG* [89], *DDX3X* [90] and *CASK* [91]).

### Widespread occurrence and clinical importance of disease “predisposing” variants

The aforementioned examples of loss-of-function variants, together with our previously discussed gain-of-function and gain of proteotoxicity *PRSS1* variants in chronic pancreatitis [19], would logically imply the widespread occurrence of “predisposing” variants in most, if not all, disease-causing genes. Our argument presupposes that mutations can potentially occur anywhere in the human genome but also that the functional impact of variants is diverse depending upon their location and type. It follows that the pathological relevance of variants in any disease-causing gene is dependent upon their functional effect. Thus, in disease-causing genes, three principal categories of variant, namely “pathogenic”, “predisposing” and “benign”, are likely to occur (Figure 5). To reinforce this concept, we cite two additional disease/gene examples. Huntington’s disease is caused predominantly by an expansion of trinucleotide CAG repeats in the huntingtin gene (*HTT*; OMIM #613004) [92]. Huntington’s disease is a true autosomal dominant condition because the clinical phenotype of a homozygote is indistinguishable from that of a heterozygote [93]. Nonetheless, it is the number of CAG repeats that determines the age of onset of disease, as well as its progression and severity. Fewer than 27 CAG repeats are asymptomatic whereas ≥ 40 CAG repeats invariably cause the disease [94]. Lying in between is an intermediate category comprising a series of CAG repeats of varying size, some of which may be classified as ‘predisposing*’*. The second example is the fragile X syndrome/*FMR1* CGG repeat expansion [95]. In unaffected individuals in the general population, the number of CGG repeats in the 5’-untranslated region of *FMR1* is fewer than 55. In patients with fragile X syndrome, however, the number of CGG repeats invariably exceeds 200. Although individuals with 55-200 CGG repeats are clinically asymptomatic, these ‘premutation carrier’ individuals are predisposed to fragile X syndrome since CGG repeats in the range 55-200 are prone to expand into the pathological range [96].

**Figure 5.**
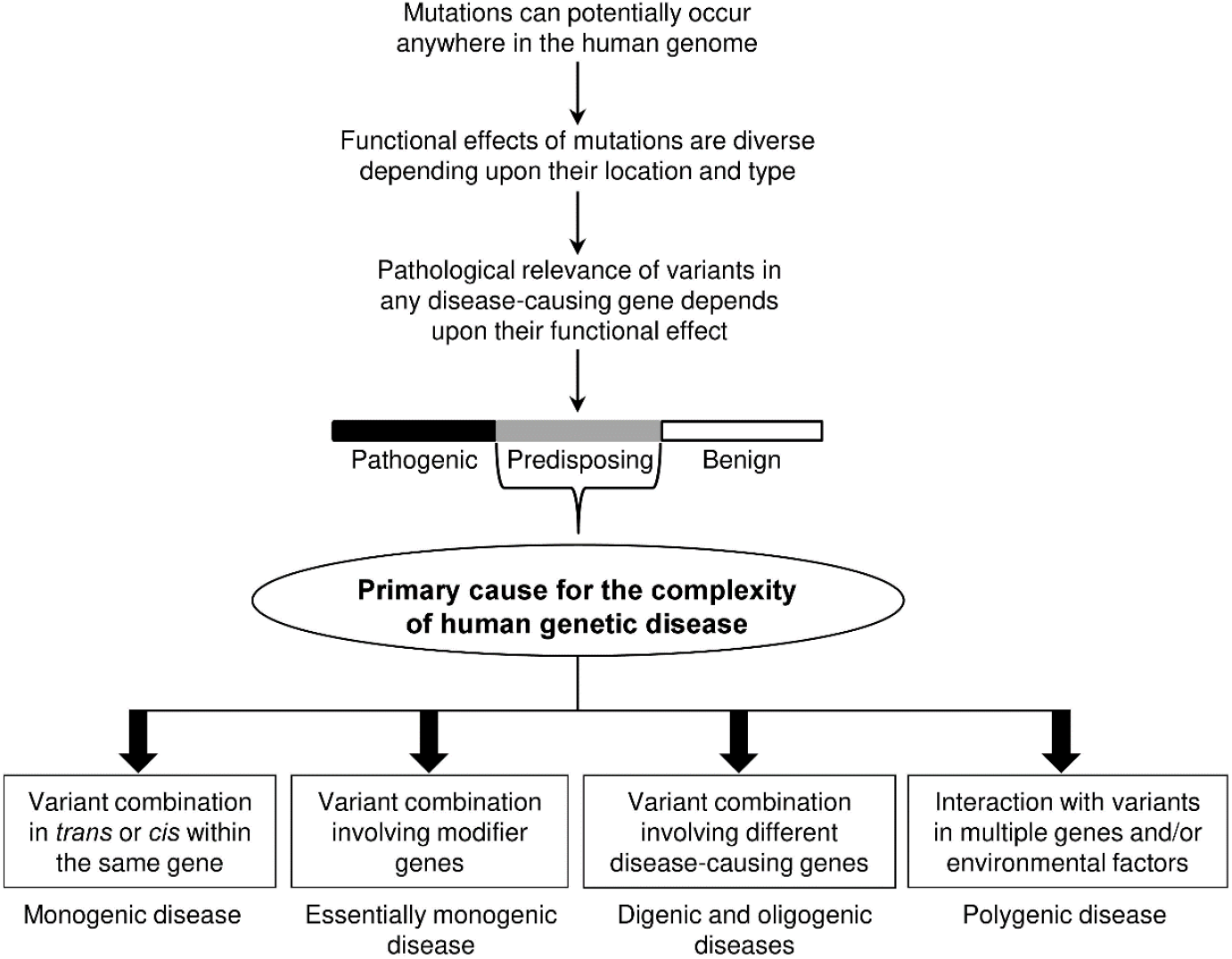
Widespread occurrence and clinical importance of “predisposing” variants. As for disease complexity, the illustrated situations should not be interpreted as representing all possible eventualities.

The importance of recognizing the new “predisposing” category, in both research and clinical contexts cannot be overstated, as “predisposing” variants are primarily responsible for the complexity of human genetic disease. In other words, the genetic basis of health and disease would be quite straightforward if all variants were either “pathogenic” or “benign”. It is difficult to list all possible contributions of “predisposing” variants to human genetic disease complexity, but some are illustrated in Figure 5. Thus, in the context of an autosomal dominant disease, the disease may appear earlier and be more severe if a “predisposing” variant occurs in *trans* or *cis* with a “pathogenic” variant in the same disease gene; further, the combination of two individually harmless “predisposing” alleles within the same gene may result in a typical or even a severe form of the disease. Moreover, a given disease may be regarded as essentially monogenic despite the involvement of modifier genes (one illustrative example might be a CF patient carrying a pathogenic *CFTR* allele, a “predisposing” *CFTR* allele and a variant in an unlinked modifier gene [97]). Further, two “predisposing” variants in independent but mechanistically linked disease-causing genes (e.g., ADPKD-causing *PKD1* and *PKD2* [48, 98]) may also be envisaged to cause a typical digenic disease. Finally, “predisposing” variants in a disease-causing gene may even contribute to disease under a polygenic inheritance or complex model (e.g., [99]).

The recognition of the new “predisposing” category will also have pivotal consequences for reproductive genetic counseling. For example, a couple each carrying a *bona fide* “predisposing” variant in an autosomal dominant disease-causing gene, could give birth to a clinically affected child. Litigation might ensue if such variants were to be inadequately and perfunctorily ascribed the descriptor “VUS” during antenatal genetic counselling.

## Discussion

Both the “hypomorphic” *PKD1* variants and the *CFTR* variants of “varying clinical consequence” serve to illustrate our dilemma in classifying clinically detected variants using the ACMG/AMP-recommended five-category scheme. We propose that this dilemma may now be resolved by adopting a new classificatory scheme with a new variant term, “predisposing”.

Based upon existing knowledge and reasonable extrapolations, we infer the presence of “predisposing” variants in most, if not all, disease-causing genes. Although this analysis focused upon loss-of-function variants, “predisposing” variants can have other functional consequences, e.g. gain-of-function or gain of proteotoxicity, depending upon the gene responsible and the underlying disease mechanism. Here, it is worth reiterating that the study of gain-of-function and gain of proteotoxicity *PRSS1* variants contributed to the proposition of our general variant classification framework [19].

Various examples of “hypomorphic” *PKD1* variants were identified through the study of exceptional families on the one hand and the availability of renal cysts as a reliable and objective diagnostic criterion for PKD on the other. Examples of *CFTR* VCC variants were identified through the careful evaluation of CF patients worldwide by the CFTR2 International Initiative. Since “predisposing” variants have by definition less profound effects than standard “pathogenic” variants, we may infer that many “predisposing” variants are likely to have been overlooked in both a research and a clinical setting. In this regard, it is pertinent to cite the examples of the abovementioned *PKD1* p.Arg3277Cys and *MECP2* p.Pro399Leu variants, both of which should theoretically have been filtered out by reference to the maximum allele frequency expected for a typical pathogenic variant in these disorders. As such, “predisposing’ variants, which can potentially include any type of variant (e.g., missense, intronic, regulatory), may not only account for a significant fraction of the unexplained cases of both monogenic and oligogenic disease [100-103] but may also be relevant to the “missing heritability” problem in complex disease [104, 105].

As illustrated in Figure 1, our new classificatory categories could be considered to be separated or spit from ACMG/AMP’s “uncertain significance” category, further facilitating the practical application of our seven-category variant classification system. Specifically, variants falling into the “predisposing” category should be predominantly present within the variants of “uncertain significance” that were classified according to the ACMG/AMP’s five-category scheme. A variant can be classified as “predisposing” if and when there is sufficient genetic and/or functional evidence to support such a classification; otherwise, one can always retain the VUS classification.

The above notwithstanding, in practice, one often has to make fairly arbitrary decisions with regard to separating “predisposing” variants from “pathogenic” and “benign” variants. This is a difficult but unavoidable task, and generally involves making arbitrary decisions with respect to the definitions of “likely pathogenic” and “likely benign” variants as well as the thresholds used for defining “high”, “moderate” or “low” penetrance. In this regard, it is pertinent to cite the “ABC” system, whose A, B and C categories all refer to “pathogenic” variants, albeit with differences in terms of their penetrance [18]. Moreover, the ClinGen Low-Penetrance/Risk Allele Working Group aims to develop a consensus with regard to the terminology required to categorize both risk alleles and low-penetrance Mendelian variants and to develop a standardized classification framework to evaluate these types of variant [106]. In our view, variants with a penetrance below a given threshold (e.g., 20%) may be more appropriately classified as “predisposing” rather than “pathogenic”. Nonetheless, given the intrinsic complexity of biology and disease, we would recommend that such decisions “need to be made on a gene-by-gene basis and would require close collaboration between researchers and clinicians with specific expertise in the diseases/genes in question” [19]. This need becomes even more pressing if we consider that (i) the same variant may be “predisposing” in the context of a typical disease but “pathogenic” in the context of a related atypical disease (e.g., *CFTR* VCC variants in CF and *CFTR*-related diseases) and (ii) variants in a given gene can potentially contribute to a disease that can manifest as monogenic, digenic/oligogenic or even multifactorial (e.g., chronic pancreatitis [19, 107]).

Over the past years, we have witnessed near continuous revolution in terms of the standards and rules governing how to weigh and combine evidence with a view to improving our classification of variants, as reflected by the abovementioned works that have attempted to adapt or refine the ACMG/AMP guidelines [2-17]. Directly or indirectly related to these adaptations or refinements, variant reclassifications (either upgrades or downgrades) have been a frequent occurrence in the literature (e.g., see recent reports [108-118]. It is our hope that our proposed general variant classification framework will stimulate discussion and debate and lead to further improvements in variant classification.

## Conclusions

In this analysis, we present cogent arguments in support of the applicability and utility of our seven-category variant classification system for disease-causing genes. The recognition of the new “predisposing” category not only has important implications for variant detection and interpretation but should also have important consequences for reproductive genetic counseling.

## Data Availability

All supporting data are available within the article.

## Authors’ contributions

J.M.C. conceived and designed the study, performed the literature search, prepared all tables and figures and drafted the manuscript. E.M., W.B.Z., Z.L., E.G., D.N.C. and C.F. contributed to the study design and critically revised the manuscript with important intellectual input. All authors approved the final manuscript submitted.

## Acknowledgements

We are grateful to Peter C. Harris (Mayo Clinic and Foundation, Rochester, Minnesota) and Karen Raraigh (Johns Hopkins University, Baltimore, Maryland) for their prompt and positive replies to our inquiries about using data from the ADPKD variant database and the CFTR2 database, respectively.

